# Evidence of depression affecting multimorbidity in survey data from the Brazilian population

**DOI:** 10.1101/2023.03.18.23287162

**Authors:** Luis Fernando Silva Castro-de-Araujo, Jacyra Azevedo Paiva de Araujo, Elisângela da Silva Rodrigues, Rodrigo Lins Rodrigues, Richard A Kanaan

**Affiliations:** Center of Data and Knowledge Integration for Health (CIDACS), Fiocruz, Salvador, Brazil; Dept of Psychiatry, The University of Melbourne, Austin Health, Victoria, Australia; Center of Data and Knowledge Integration for Health (CIDACS), Fiocruz, R. Mundo, 121. Salvador, Bahia, Brazil; Federal University of Ceará, Campus Jardins de Anita, Itapajé, Ceará, Brazil; Rural Federal University of Pernambuco, Department of Education (DED), Recife, Pernambuco, Brazil; Department of Psychiatry, University of Melbourne, Austin Health, Heidelberg, VIC 3084, Australia

**Keywords:** multimorbidity, depression, structural equation modeling

## Abstract

**Objectives:** Depression is associated with multimorbidity, the occurrence of two or more chronic diseases. Although the effect of multimorbidity on depression is relatively well known, the opposite effect is less well studied. We aimed to examine the effect of depression on multimorbidity using data from a nationally representative Brazilian survey.

**Methods:** We used information from all respondents above 15 years of age of the Brazilian National Survey on Access, Use, and Promotion of the Rational Use of Medicines (PNAUM) from 2014. A structural equation model was fit to the data with a specification that included the relationship between depressive syndrome and multimorbidity, controlled by age and body mass index.

**Results:** The data set comprised 28,382 subjects. The model presented fit the data well, and revealed a statistically significant, positive, moderate-size effect (0.355) of depression on multimorbidity.

**Conclusion:** Depression appears to make a moderate contribution to the development of multimorbidity.

## Introduction

Depression is one of the leading health concerns worldwide today^1,2^. Over 350 million people are affected by this disorder and prevalence in the general population is estimated between 3 to 10%^1^. It is the single largest cause of years lost to disability^2^, as it is a chronic disorder with symptoms that last for long periods and are recurring. It is also related to worse quality of life, medical morbidity and higher mortality^3^.

There is evidence that depression is associated with multimorbidity, the occurrence of two or more chronic diseases. It has been shown that the direction of this association seems to run from multimorbidity to depression^4,5^. As individuals age, they tend to become alone, losing friends and family members, which renders them susceptible to depressive symptoms^6^. Furthermore, with age they are subject to brain vascular injury due to hypertension, diabetes mellitus, obesity, and high blood cholesterol^7^. Examining the relation between multimorbidity and depression is important as a global health preventive measure, as it can help match resources to the individuals who are hit the hardest in crises like the recent Covid19 pandemic. For example, during the COVID19 pandemic, multimorbidity was associated with increased anxiety and depressive symptoms both during the social isolation phase ^8^ and post-infection^9^. A systematic review published by Read et al. (2017) reported that people with multimorbidity have twice the risk of depressive disorders^7^. Socioeconomic factors seem to play an important role in the multimorbidity of mental disorders and chronic diseases, as this combination of disorders are more common in more deprived areas, where they particularly affect younger individuals^10,11^.

Multimorbidity is known to increase with age, reaching higher prevalence in very old individuals. There is an ongoing effort of trying to identify patterns of clustering for these conditions, which varies significantly across studies^12^. A three-cluster pattern seems to be emerging, with musculoskeletal, cardiometabolic, and neuropsychiatric clusters^4,12^. Furthermore, being overweight is associated with multimorbidity in a dose-response pattern. A recent Finnish study reported a hazard ratio of 2.83 for obesity and 2.67 for those overweight^13^ in developing multimorbidity. It is therefore essential to take ageing and body mass index (BMI) into consideration in every multimorbidity study.

By contrast, the effect of depression on multimorbidity is seldom investigated. Subjects with depression are vulnerable to multimorbidity due to poor lifestyle choices (such as tobacco smoking or taking less exercise), limited adherence to clinical treatment and poor access to health^14^. There is also evidence that depression and multimorbidity have common biological and psychosocial drivers (inflamation, stress, economic burden) that might interact and result in one or both conditions^18^. Additionally, some medications for depression are associated with metabolic syndromes, obesity, and cardiovascular events^15–17^. So, though it is clear that multimorbidity has an important effect on depression, there is a potential causal path in the opposite direction. This paper aims to investigate this, the effect of depression on multimorbidity, using a publicly available Brazilian survey including 28,382 respondents from 2014, and incorporating age and BMI as controls, in a structural equation model that accounts for error measurement.

## Methods

### Data source

Data from the National Survey on Access, Use, and Promotion of the Rational Use of Medicines in Brazil (PNAUM) were used in this study. The interviews for this survey took place between September 2013 and February 2014^14^, and were carried out by 165 trained interviewers. Only permanent residents of urban areas of Brazil were interviewed. All respondents in the available set were included in our analysis, which ranged from 15 to 99 years. Data were downloaded from the PNAUM website (http://www.ufrgs.br/pnaum/documentos/micro-dados/microdados-adultos/view) and pre-processed by RLR to allow analysis in the R statistical environment. The PNAUM data collection was described previously ^19^. The data is public domain and is freely available from the website, along with raking weights for each respondent. The raking was performed according to the population distribution obtained in the 2013 Brazilian National Survey on Health by age, gender and region. The 2013 Brazilian National Survey (PNS 2013) is a Survey carried out by the Brazilian Institute of Geography and Statistics to evaluate the performance of the National Health System in terms of access and use of available services, as well as the surveillance of the chronic non-communicable diseases and related risk factors (https://www.ibge.gov.br/estatisticas/sociais/saude/29540-2013-pesquisa-nacional-de-saude.html?=&t=microdados).

### Variables

The included variables were: age; body mass index (BMI); age when respondent was diagnosed with depression; psychiatric medication use, whether the respondent was using any psychiatric medication at the time of the interview; number of medications for chronic clinical diseases; cumulative number of chronic clinical diseases; multimorbidity, whether the patient had multimorbidity or not coded as 0/1 and included in the model as continuous; time medicated, time in years since beginning to use medications for chronic clinical conditions. In the SEM specification, the variables for depression severity, psychiatric medication use, and time since depression diagnosis were used to specify a reflective latent variable representing depression syndrome. The chronic-disease related variables were used to specify a reflective latent variable representing multimorbidity. One particular question was not standard and was not validated in previous studies, the depression severity question. The interviewers asked “how much limitation was due to the respondent diagnosis of depression”, without specifying the type of limitation. It was coded on a likert scale (1, no limitations; 2, mildly; 3, moderately;4, severe;5, very severe). In the pre-processing of this data, knowing the limitations of the statistical package we used, we re-coded this variable so that respondents that did not have depression would be coded as zero.

### Data analysis

All analyses were performed using R version 4.1.3 (https://cran.r-project.org/). All analyses used an alpha of 0.05. The overall missingness of the entire dataset was below 10% (Supplemental material). Missing data were filled by pooling from 5 multiply-imputed data frames using the R mice package with the classification and regression trees algorithm^20^. The analytic strategy used structural equation modelling, allowing both investigation of latent constructs and accounting for measurement error. The structural equation model specification can be seen in Figure 1. The estimation of the coefficients was performed using the lavaan survey package version 1.1.3.1 and incorporated the rake weights. This package does not allow categorical variables. This is the reason all variables were included in the model as continuous. For complex survey data, i.e. with weights, there currently is no alternative to this limitation. Identification was automatically tested by loading the model specification into the umx package^21^ and using the mxCheckIdentification tool^22^. The model is locally identified. The path diagram of figure 1 was built using Onyx^23^. The estimation was maximum likelihood based, using the “MLM” option to obtain robust fit statistics.

**Figure 1.**
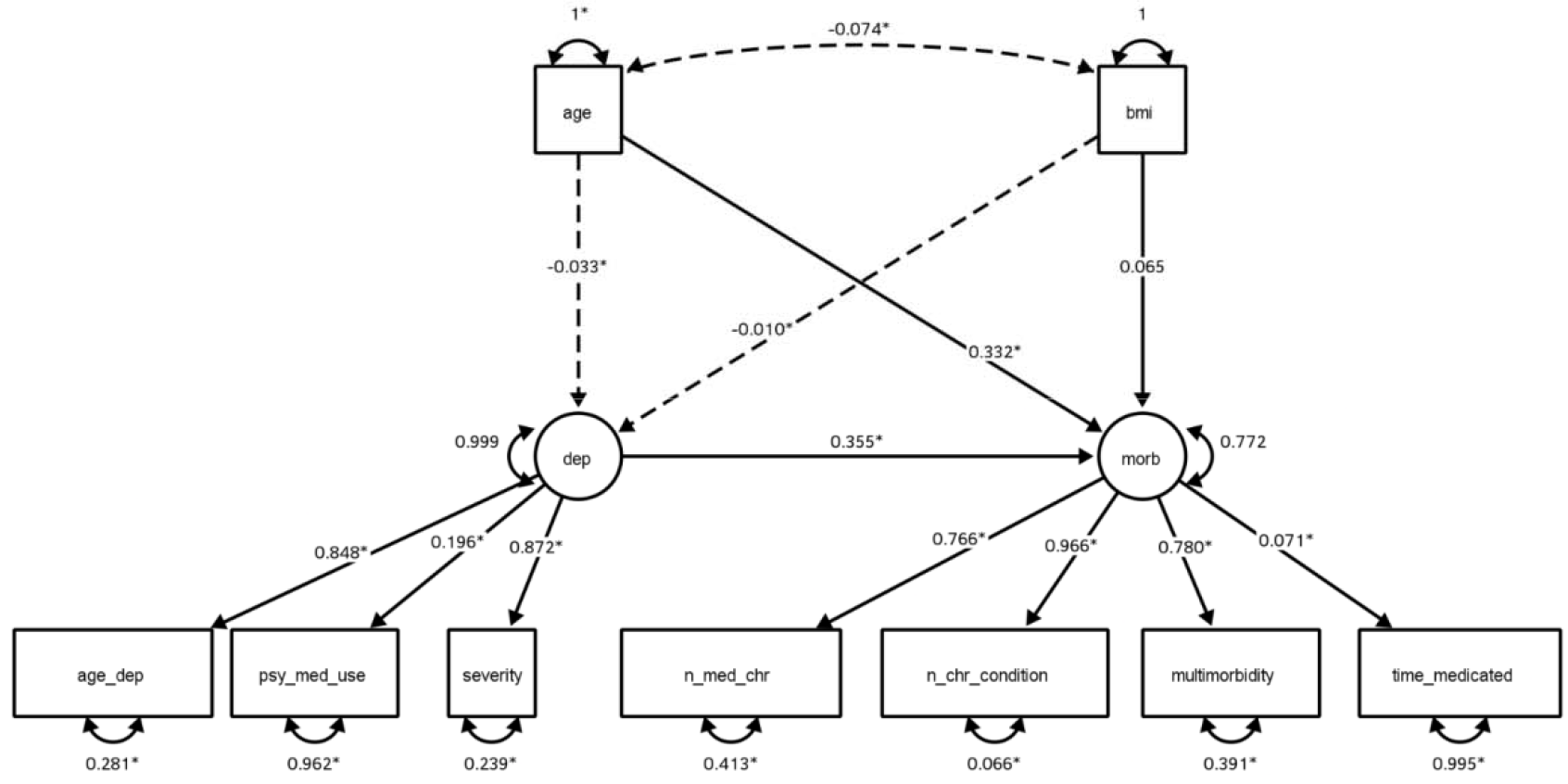
SEM path specification. Dashed lines are negative coefficients. Standard SEM representation is used, circles are latent variables, boxes are measured variables. Standardized estimates. Model Fit: χ^2^(23) = 671.442, p < 0.001; Robust CFI = 0.971;Robust TLI = 0.955; Robust RMSEA = 0.051 (90%CI0.048,0.055); Robust SRMR = 0.026. *, statistically significant. Age_dep, age in which the respondent was diagnosed with depression; Psy_med_use, whether the respondent used psychiatric medications; Severity, variable coding whether the respondent had depression and if yew, the burden it produced in their lives (no, mild, severe, very severe); n_med_chr, number of medications used to treat chronic conditions; n_chr_condition, number of chronic conditions; multimorbidity, whether the respondent endorsed multimorbidity; time_medicated, for how long the respondent has been using medications

## Results

After pre-processing, the set contained 28,382 respondents. Demographic and clinical statistics are shown in Table 1. Continuous variables were analysed with the average of the descriptive measures, median, minimum, maximum and standard deviation (SD) and categorical variables were described with the total number of observations and their corresponding percentages stratified by sex. The data set consisted of 8,780 (30.9%) males and 19,602 (69.1%) females. The individuals had a mean age of 61.04 (SD 14.04) years, and most patients were aged between 60 and 99 years (Males: 60.4% and Females: 52.7%). Both males and females had on average 2 chronic diseases (Males: 2.05 (SD 1.30) and Females: 2.30 (SD 1.38)). This was also observed in the number of medications for chronic disease treatment (Males: 2.10 (SD 1.28) and Females: 2.30 (SD 1.32)). Most individuals resided in the South (Males: 25.3% and Females: 25.0%) and Southeast regions (Males: 23.8% and Females: 23.6%), while individuals from the North of the country were the minority in this data set (Men: 14.9% and Women : 12.9%). Of those responding, 64.8% were considered multimorbid, 59.6% of males and 67.2% of females. Only 0.8% of respondents used psychiatric medication, 0.4% of males and 1.1% of females. Females engaged in intense physical exercise on average 7.06 (SD 22.25) days per week, while males exercised on average 4.62 (SD 10.26) days per week. Males smoked on average 17.30 (SD 18.44) cigarettes per day and females 15.11 (SD 17.36) cigarettes per day. On average, individuals with depression were diagnosed at 48.48 (SD 21.02) years of age; males at age 52.12 (SD 19.74), and females at age 47.76 (SD 21.19). In response to the depression severity variable, the majority of individuals did not endorse any life limitations from depression (Males: 4.3% and Females: 8.4%) others acknowledged few limitations (Males: 3.0% and Females: 7.2%), moderate limitations (Males: 1.7% and Females: 4.1%), intense limitations (Males: 1.1% and Females: 2.9%) and very intense limitations (Males: 0.7% and Females: 1.6%). Note that this variable also encoded subjects who did not have a diagnosed depression, which was the majority of both sexes (Table 1).

**Table 1.**
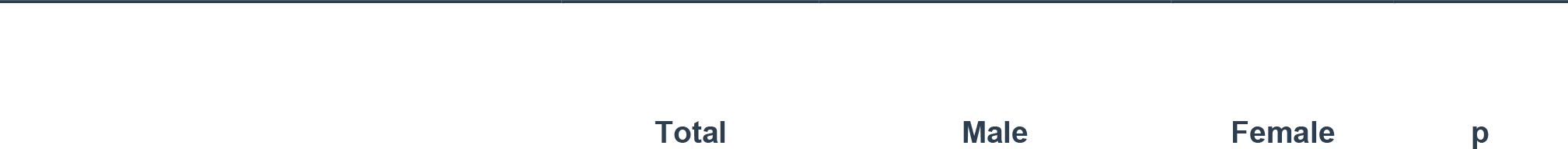
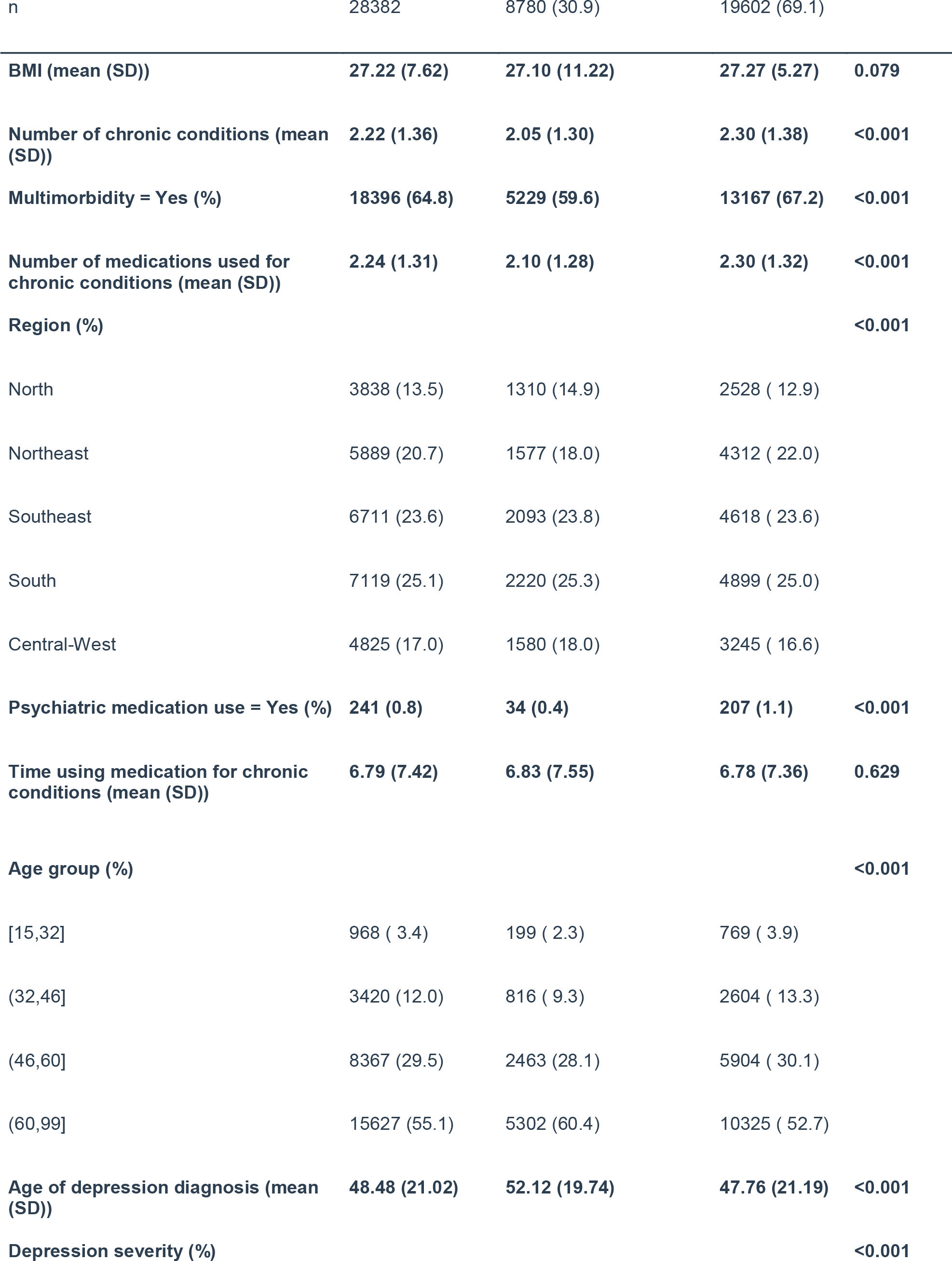
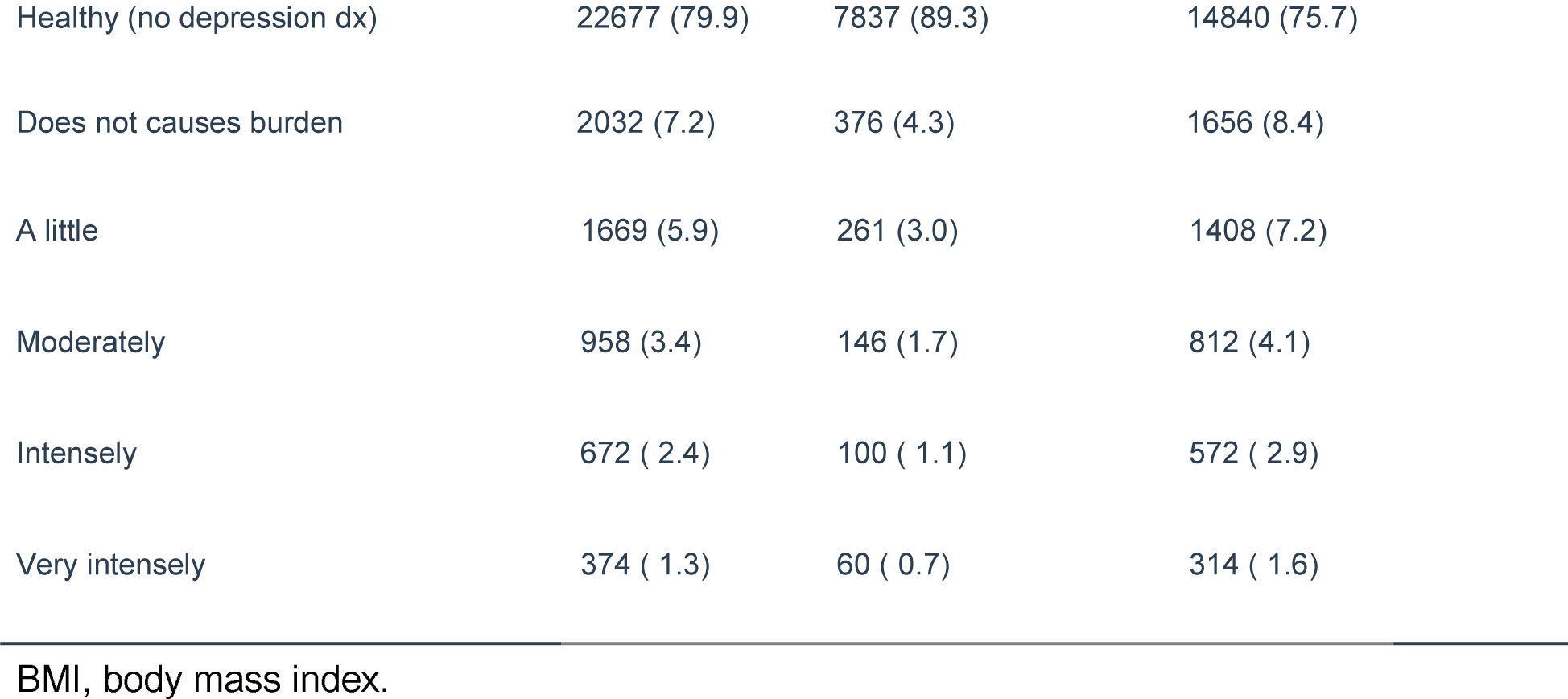
Clinical and demographic data stratified by sex. Group comparison was performed using Chi-2 contingency table tests for the categorical variables and ANOVA for the continuous variables.

| Total | Male | Female | p |
| --- | --- | --- | --- |

|  |  |  |  |  |
| --- | --- | --- | --- | --- |
| n | 28382 | 8780 (30.9) | 19602 (69.1) |  |
| **BMI (mean (SD))** | **27.22 (7.62)** | **27.10 (11.22)** | **27.27 (5.27)** | **0.079** |
| **Number of chronic conditions (mean (SD))** | **2.22 (1.36)** | **2.05 (1.30)** | **2.30 (1.38)** | **<0.001** |
| **Multimorbidity = Yes (%)** | **18396 (64.8)** | **5229 (59.6)** | **13167 (67.2)** | **<0.001** |
| **Number of medications used for chronic conditions (mean (SD))** | **2.24 (1.31)** | **2.10 (1.28)** | **2.30 (1.32)** | **<0.001** |
| **Region (%)** |  |  |  | **<0.001** |
| North | 3838 (13.5) | 1310 (14.9) | 2528 ( 12.9) |  |
| Northeast | 5889 (20.7) | 1577 (18.0) | 4312 ( 22.0) |  |
| Southeast | 6711 (23.6) | 2093 (23.8) | 4618 ( 23.6) |  |
| South | 7119 (25.1) | 2220 (25.3) | 4899 ( 25.0) |  |
| Central-West | 4825 (17.0) | 1580 (18.0) | 3245 ( 16.6) |  |
| **Psychiatric medication use = Yes (%)** | **241 (0.8)** | **34 (0.4)** | **207 (1.1)** | **<0.001** |
| **Time using medication for chronic conditions (mean (SD))** | **6.79 (7.42)** | **6.83 (7.55)** | **6.78 (7.36)** | **0.629** |
| **Age group (%)** |  |  |  | **<0.001** |
| [15,32] | 968 ( 3.4) | 199 ( 2.3) | 769 ( 3.9) |  |
| (32,46] | 3420 (12.0) | 816 ( 9.3) | 2604 ( 13.3) |  |
| (46,60] | 8367 (29.5) | 2463 (28.1) | 5904 ( 30.1) |  |
| (60,99] | 15627 (55.1) | 5302 (60.4) | 10325 ( 52.7) |  |
| **Age of depression diagnosis (mean (SD))** | **48.48 (21.02)** | **52.12 (19.74)** | **47.76 (21.19)** | **<0.001** |
| **Depression severity (%)** |  |  |  | **<0.001** |
| Healthy (no depression dx) | 22677 (79.9) | 7837 (89.3) | 14840 (75.7) |
| Does not causes burden | 2032 (7.2) | 376 (4.3) | 1656 (8.4) |
| A little | 1669 (5.9) | 261 (3.0) | 1408 (7.2) |
| Moderately | 958 (3.4) | 146 (1.7) | 812 (4.1) |
| Intensely | 672 ( 2.4) | 100 ( 1.1) | 572 ( 2.9) |
| Very intensely | 374 ( 1.3) | 60 ( 0.7) | 314 ( 1.6) |

BMI, body mass index.

The path of interest for the SEM was the regression between the latent variable representing depression to the latent variable representing multimorbidity. The model fit was good: χ^2^(23) = 671.442, p < 0.001; Robust CFI = 0.971; Robust TLI = 0.955; Robust RMSEA = 0.051 (90%CI 0.048,0.055); Robust SRMR = 0.026. All observed variables were modelled with error measurement variances (Figure 1). The standardized coefficient found for this path was 0.355 and was statistically significant (p<0.05), which means that for each SD increase in the depression syndrome latent variable, an increase of 0.35 SD in multimorbidity is expected. This effect is medium size. Age contributed a positivemoderate effect on multimorbidity, and BMI contributed a small positive effect on multimorbidity, though neither were statistically significant). The upper part of the model (BMI and age) was specified so that these variables could act as controls in the relationship between depression and multimorbidity.

Age and BMI both had significant small negative effects on the depressive syndrome latent variable. The former was surprising, given that depression is positively associated with ageing^24^.

## Discussion

This is a cross-sectional, maximum likelihood based analysis of the association between depression and multimorbidity using complex survey data from Brazilian individuals. It found a significant association in the direction of depression affecting multimorbidity, a direction seldom studied in the multimorbidity literature. The size of the effect found (0.355) was medium, and the model presented a good fit in several fit indices.

Most of the present literature in multimorbidity has focused on the effect of multimorbidity in depression. We have shown that the effect of depression on multimorbidity is of importance. In our analysis, age had a moderate effect on multimorbidity (0.332) and BMI had a small effect on (0.06, not significant) multimorbidity. The former confirms the pattern found in the literature ^12^, the latter was less than what was expected based on what had been reported of the effect of BMI on multimorbidity ^13^. In this analysis, BMI and age had little effect on depressive syndrome, which was surprising give that data comprises older subjects and the known relationship between age and depression ^25^.

Obesity (measured by BMI or waist circumference) is a highly prevalent health condition associated with early death and high health costs^26^. The rising incidence of obesity in the low-middle income countries is associated with multimorbidity. Romano et al. (2021) reported based on a sample of elderly (over 60 years) individuals that obesity (BMI≥ 30 kg/m2) was associated with 1.43 times higher odds for multimorbidity. The relation was significantly positive between obesity and hypertension, arthritis, asthma, diabetes and chronic back pain^27^. In another study from Finland using participants from the UK, researchers found that individuals with obesity started presenting complex multimorbidity by age 55, whereas individuals with healthy weight would only show multimorbidity by age 75. Also, they found that the degree of obesity was associated with complex multimorbidity in a dose-response relationship^13^.

The co-occurrence of depression and multimorbidity has been described by numerous previous studies^18,28^. Zhang et al. (2022) found that multimorbidity was associated with increased odds of having depressive symptoms (adjusted odds ratio of 1.51), and developing depressive symptoms (adjusted odds ratio of 1.65). Depression worsens the outcomes of people living with multimorbidity increasing disability, accelerating cognitive decay and also increasing costs in older individuals^5,7,29^. Moreover, depression was associated with a higher incidence of somatic disease, cardiometabolic disease, musculoskeletal diseases and digestive diseases in a 6-year follow-up cohort in the Netherlands^30^. Momen et al. (2020) conducted a cohort study using 16 years of health data in Denmark and found that individuals with a mood disorder had a cumulative incidence within 15 years after the diagnosis of a mental disorder for circulatory conditions of 40.9% (95% CI, 40.5 to 41.3). Another study using the Center for Epidemiological Studies Depression Scale (CES-D) found a positive association between depression score and chronic disease incidence. Every one point symptom increase in the CES-D scale resulted in a 5% increase in incidence of chronic diseases^31^. Given the prevalence of depression, the public health implications of this are substantial.

The study findings should be interpreted in the context of some limitations. We used the term ‘effects’ throughout the paper, as is the standard in SEM analyses, but it was a cross-sectional study, and therefore should not be considered a causal investigation. In the survey used in the study, the diagnosis of depression was self-reported, and no diagnostic instrument was used. The negative coefficients from age and BMI to depression may be related to the continuous treatment of some variables that are categorical or ordinal, which might have interfered in the variances for the latent variable representing depression. Despite these limitations, SEM allowed us to model measurement error, which is expected in survey data. This is often overlooked in psychiatry research and is an important shortcoming in survey data of psychiatric outcomes. Also, the fit indices suggest that the chosen model was parsimonious.

## Conclusion

We have presented here evidence of the effect of depression on multimorbidity in a large, nationwide representative data set. Several mechanisms can explain the effect in this direction. Subjects with depression might adhere less to clinical treatments, engage less in healthy life style, exercise less often, or anti-depressant use may increase multimorbidity by its metabolic effects. However, this can only be confirmed in studies with longitudinal designs. A future direction in this field could be to investigate whether the effect of depression on multimorbidity is bidirectional, which can be assessed in quasi-experimental methods such as cross-lagged panel data models.

## Data Availability

All data produced are available online at http://www.ufrgs.br/pnaum/documentos/micro-dados/microdados-adultos/view

## Individual contributions

All authors contributed to the study conception and design. Material preparation, data collection and analysis were performed by Elisângela da Silva Rodrigues and Rodrigo Lins Rodrigues. The first draft of the manuscript was written by Jacyra Paiva de Araujo and Luis Castro-de-Araujo. Richard AA Kanaan contributed to later versions of the manuscript. All authors commented on versions of the manuscript, read and approved the final manuscript.

## Acknowledgments

LFSCA, ESR, and JAPA were funded by the Medical Research Council - UK, Grant no. MR/T03355X/1 during the study.

## Supplementary Material

### Distribution of age by sex

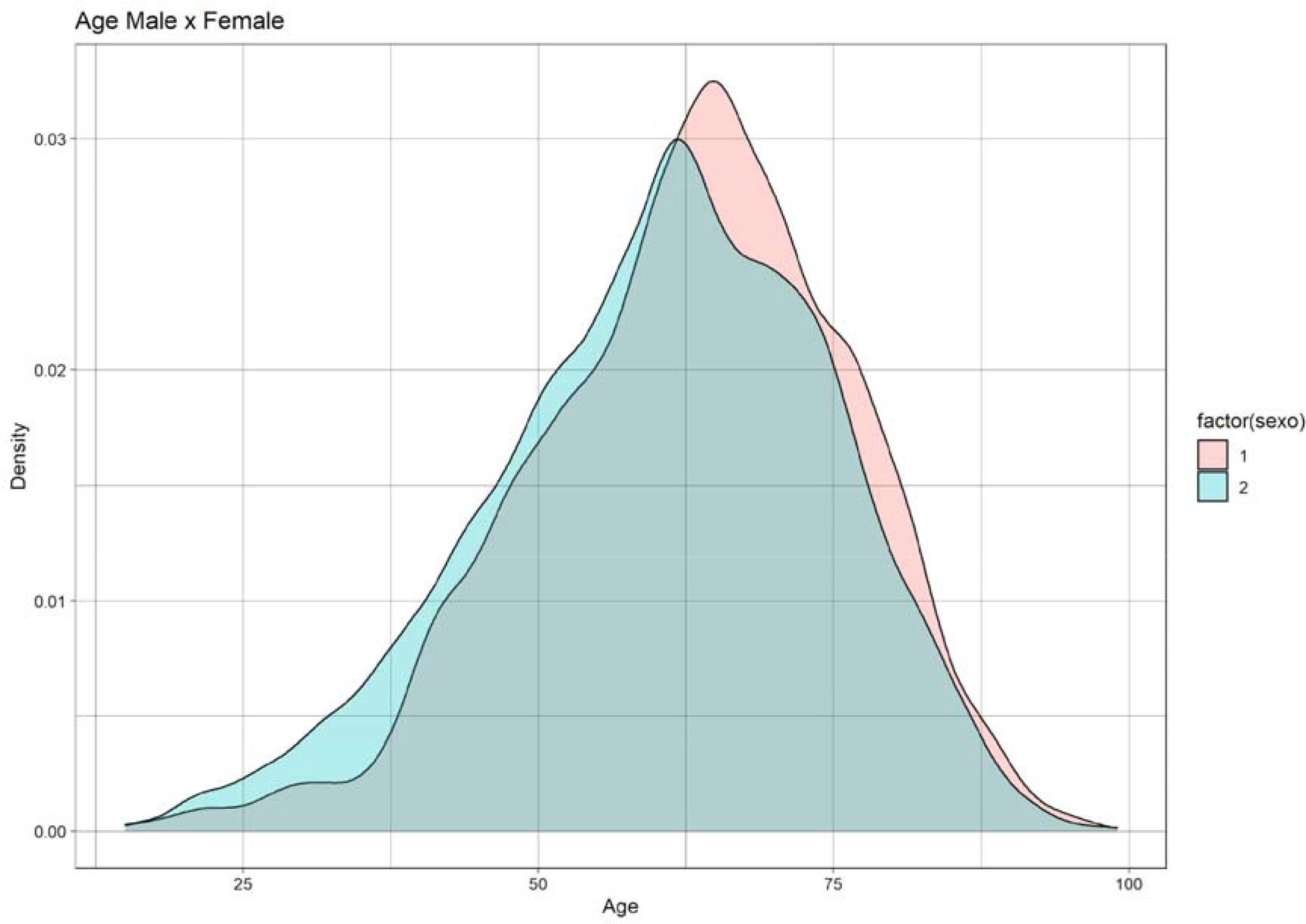

Missing and categories and distribution in one picture

*Missings, categories and distributions*

### Survey weights in structural equation modeling (SEM) with imputed data

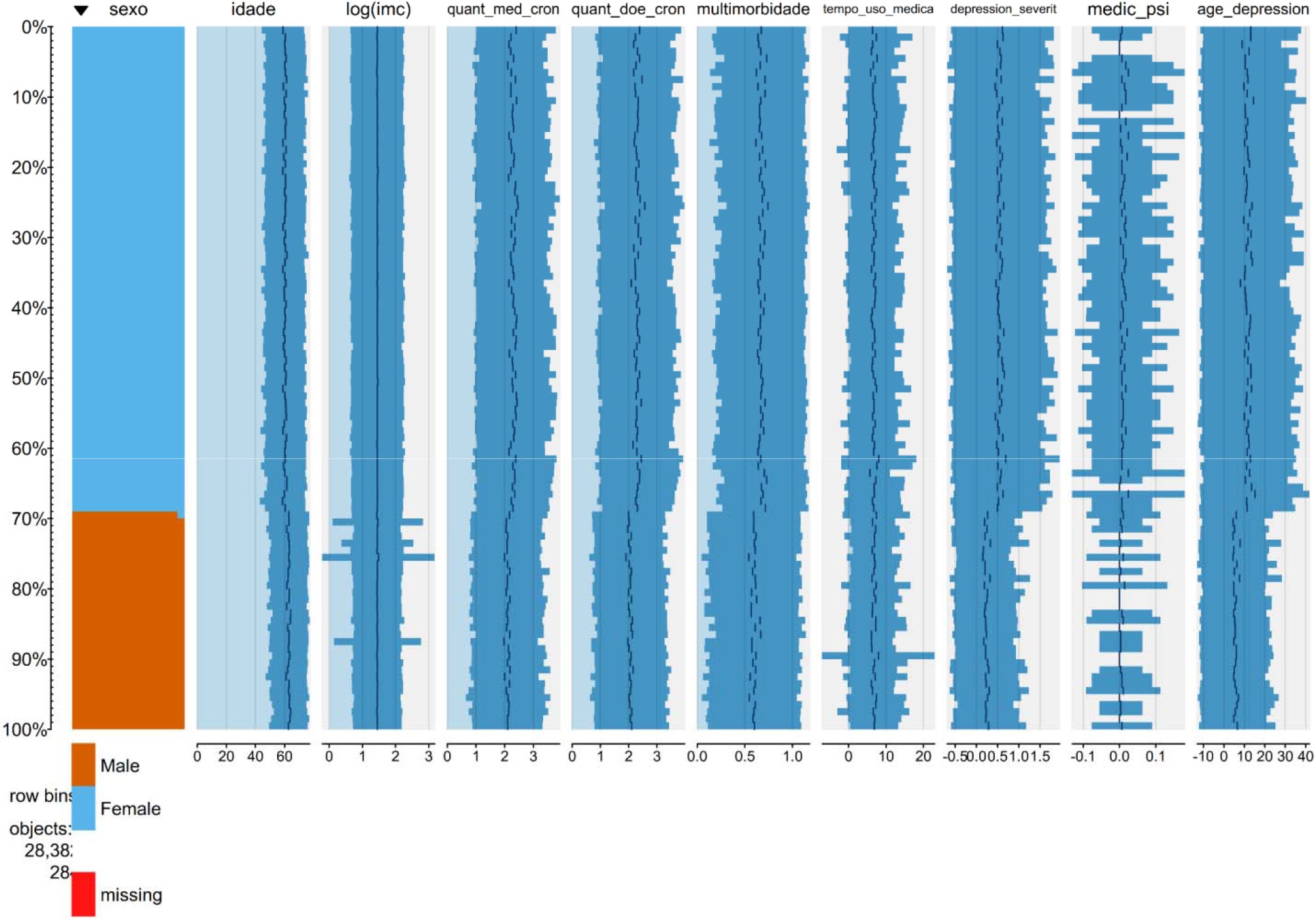

### Fit the model using weighted data (lavaan.survey.fit)

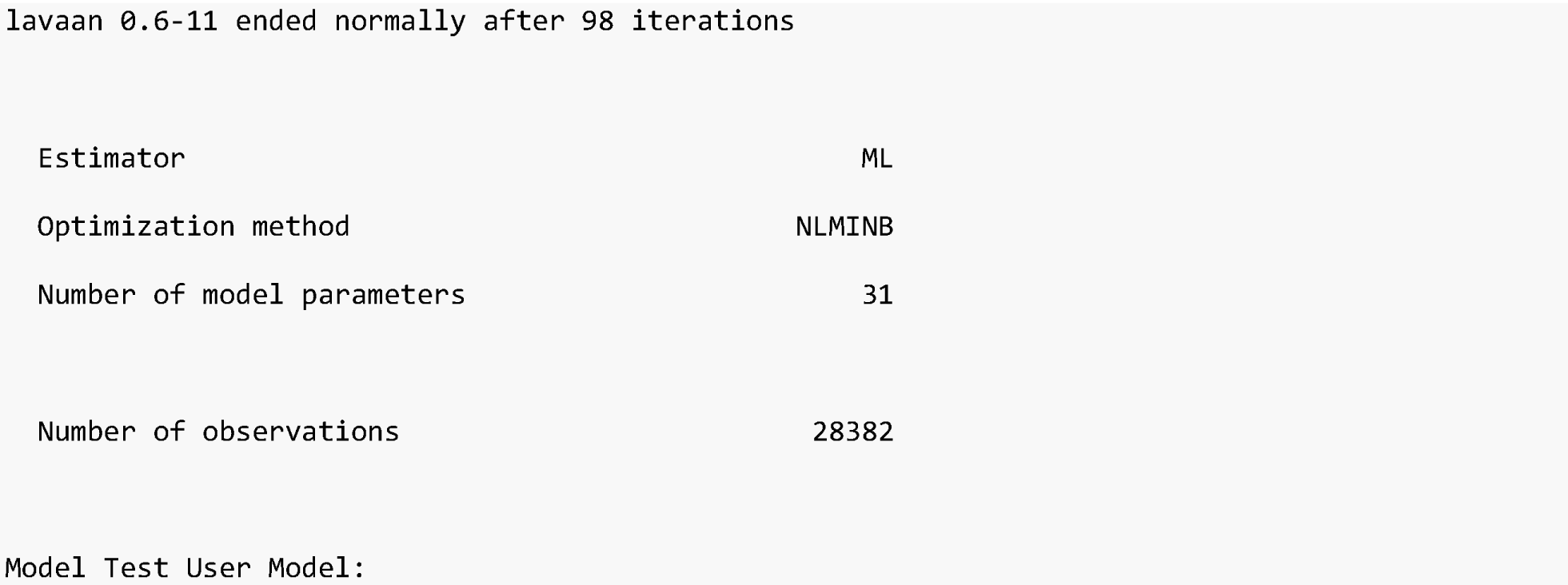
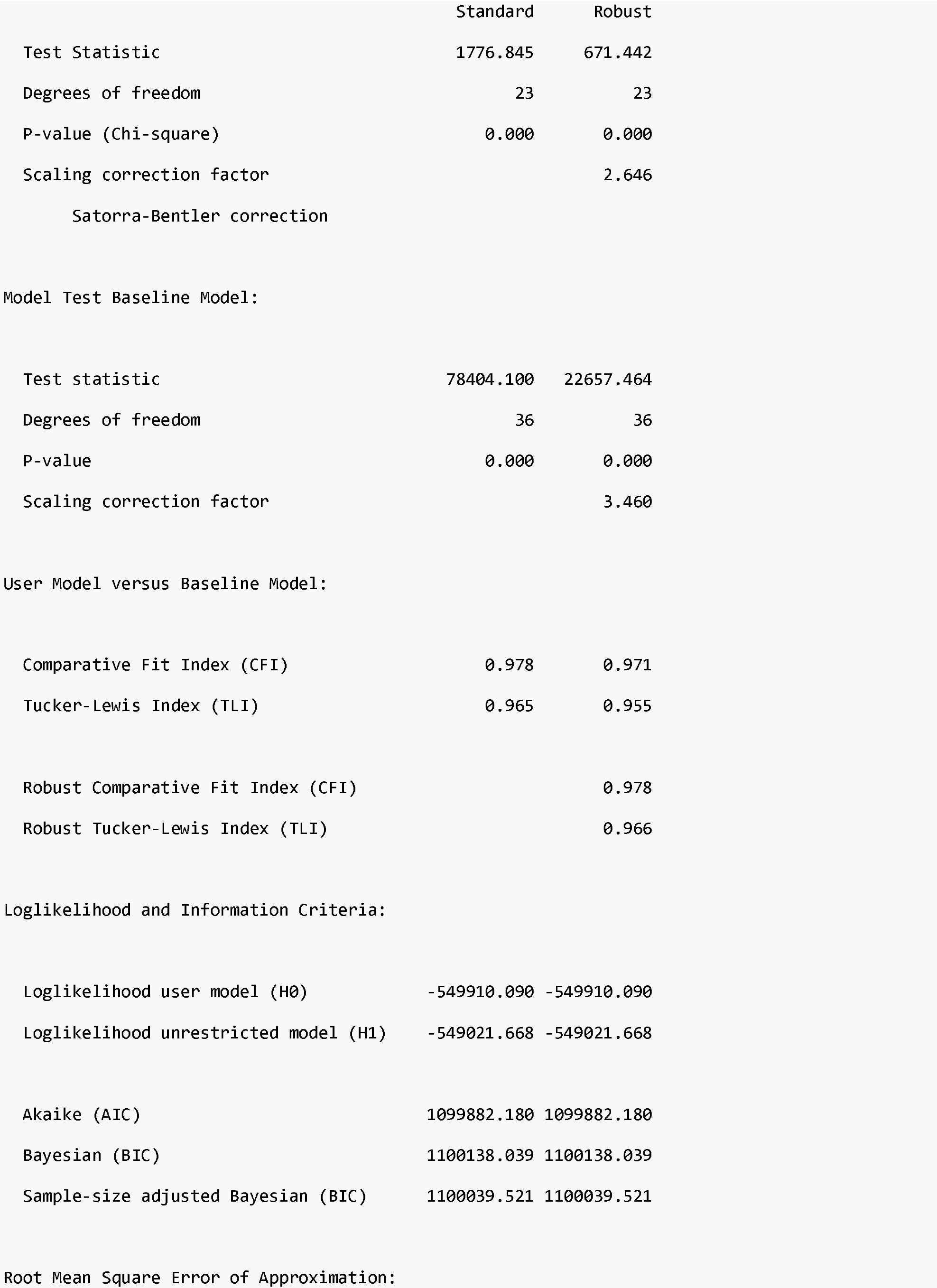
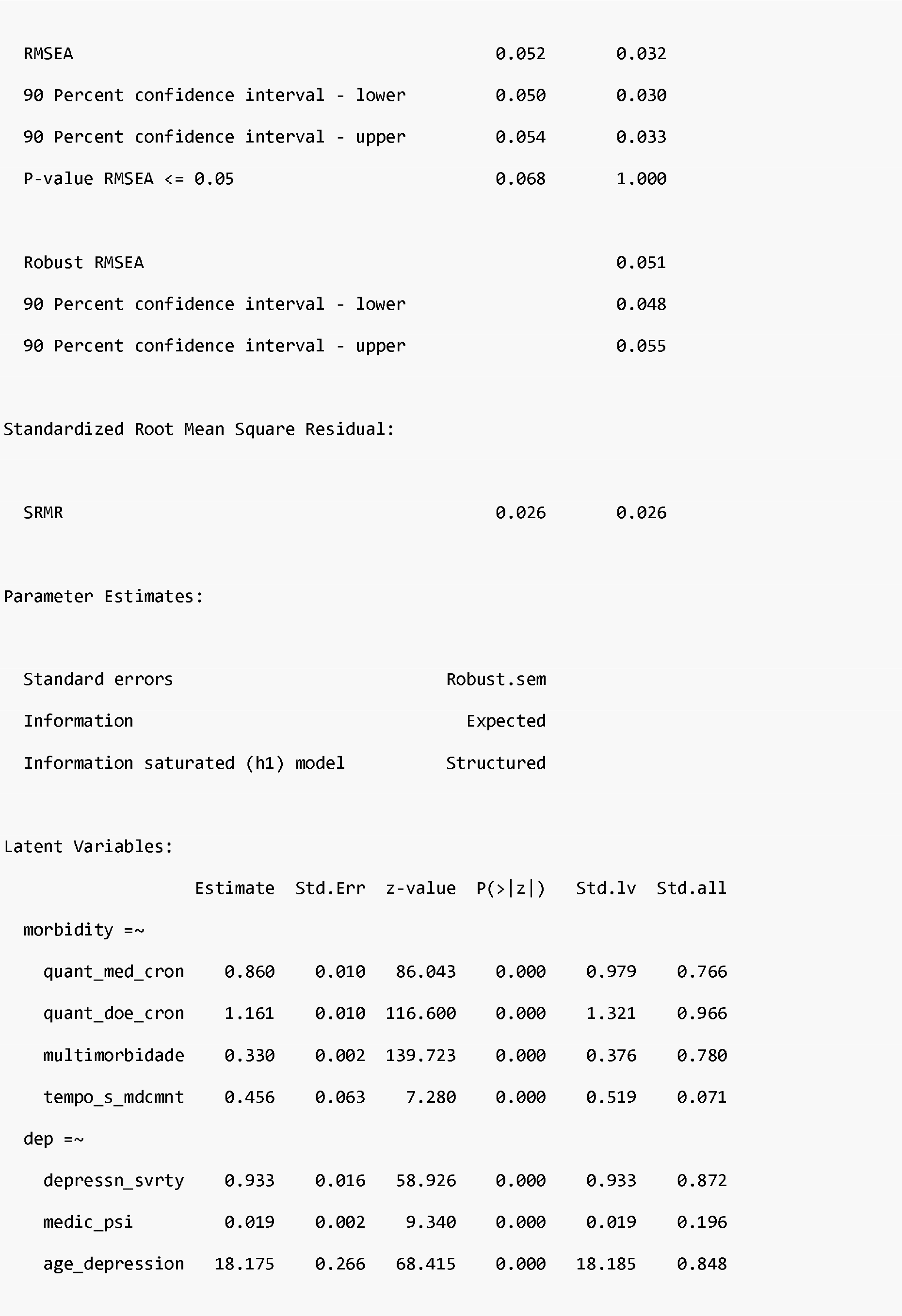
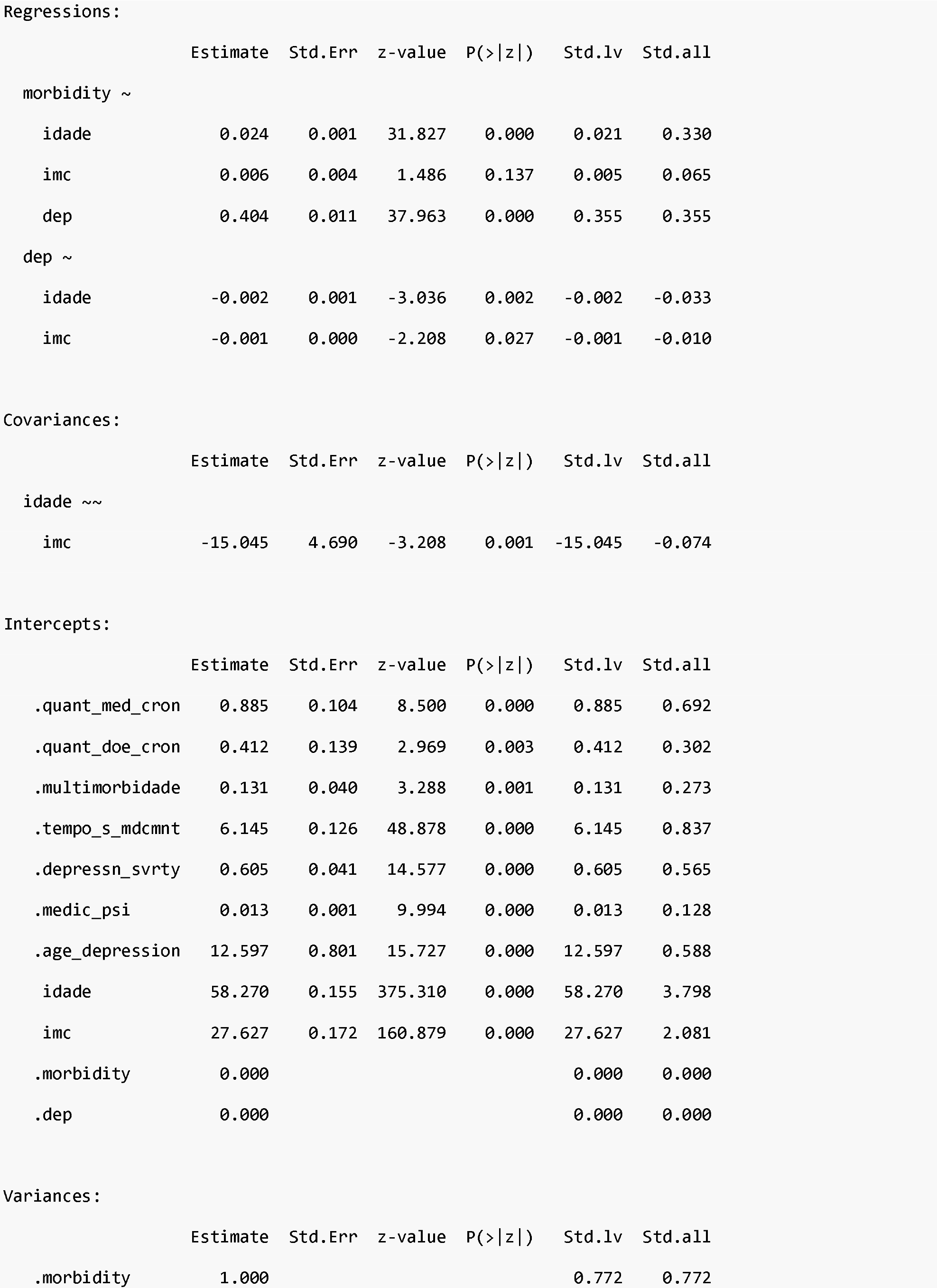
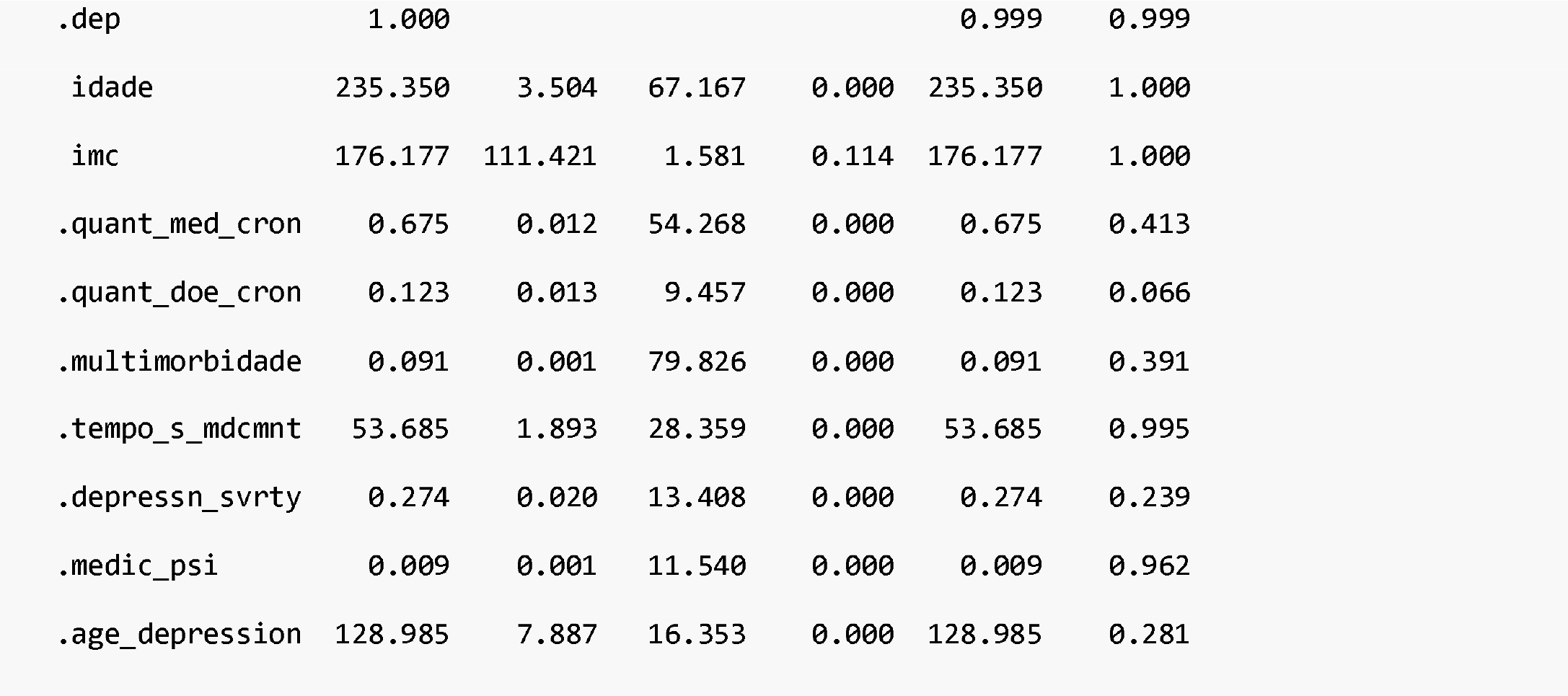

|  |  |
| --- | --- |
| Estimator | ML |
| Optimization method | NLMINB |
| Number of model parameters | 31 |
| Number of observations | 28382 |

Evidence of depression affecting multimorbidity
|  | Standard | Robust |
| --- | --- | --- |
| Test Statistic | 1776.845 | 671.442 |
| Degrees of freedom | 23 | 23 |
| P-value (Chi-square) | 0.000 | 0.000 |
| Scaling correction factor |  | 2.646 |
| Satorra-Bentler correction |  |  |
| Model Test Baseline Model: |  |  |
| Test statistic | 78404.100 | 22657.464 |
| Degrees of freedom | 36 | 36 |
| P-value | 0.000 | 0.000 |
| Scaling correction factor |  | 3.460 |
| User Model versus Baseline Model: |  |  |
| Comparative Fit Index (CFI) | 0.978 | 0.971 |
| Tucker-Lewis Index (TLI) | 0.965 | 0.955 |
| Robust Comparative Fit Index (CFI) |  | 0.978 |
| Robust Tucker-Lewis Index (TLI) |  | 0.966 |
| Loglikelihood and Information Criteria: |  |  |
| Loglikelihood user model (H0) | -549910.090 | -549910.090 |
| Loglikelihood unrestricted model (H1) | -549021.668 | -549021.668 |
| Akaike (AIC) | 1099882.180 | 1099882.180 |
| Bayesian (BIC) | 1100138.039 | 1100138.039 |
| Sample-size adjusted Bayesian (BIC) | 1100039.521 | 1100039.521 |
| Root Mean Square Error of Approximation: |  |  |

|  |  |  |
| --- | --- | --- |
| RMSEA | 0.052 | 0.032 |
| 90 Percent confidence interval - lower | 0.050 | 0.030 |
| 90 Percent confidence interval - upper | 0.054 | 0.033 |
| P-value RMSEA <= 0.05 | 0.068 | 1.000 |
| Robust RMSEA |  | 0.051 |
| 90 Percent confidence interval - lower |  | 0.048 |
| 90 Percent confidence interval - upper |  | 0.055 |

Standardized Root Mean Square Residual:
|  |  |  |
| --- | --- | --- |
| SRMR | 0.026 | 0.026 |
| Parameter Estimates: |  |
| Standard errors | Robust.sem |
| Information | Expected |
| Information saturated (h1) model | Structured |

Latent Variables:
|  | Estimate | Std.Err | z-value | P(>\|z\|) | Std.lv | Std.all |
| --- | --- | --- | --- | --- | --- | --- |
| morbidity =~ |  |  |  |  |  |  |
| quant_med_cron | 0.860 | 0.010 | 86.043 | 0.000 | 0.979 | 0.766 |
| quant_doe_cron | 1.161 | 0.010 | 116.600 | 0.000 | 1.321 | 0.966 |
| multimorbidade | 0.330 | 0.002 | 139.723 | 0.000 | 0.376 | 0.780 |
| tempo_s_mdcmnt | 0.456 | 0.063 | 7.280 | 0.000 | 0.519 | 0.071 |
| dep =~ |  |  |  |  |  |  |
| depressn_svrty | 0.933 | 0.016 | 58.926 | 0.000 | 0.933 | 0.872 |
| medic_psi | 0.019 | 0.002 | 9.340 | 0.000 | 0.019 | 0.196 |
| age_depression | 18.175 | 0.266 | 68.415 | 0.000 | 18.185 | 0.848 |

Evidence of depression affecting multimorbidity Regressions:
|  | Estimate | Std.Err | z-value | P(>\|z\|) | Std.lv | Std.all |
| --- | --- | --- | --- | --- | --- | --- |
| morbidity ~ |  |  |  |  |  |  |
| idade | 0.024 | 0.001 | 31.827 | 0.000 | 0.021 | 0.330 |
| imc | 0.006 | 0.004 | 1.486 | 0.137 | 0.005 | 0.065 |
| dep | 0.404 | 0.011 | 37.963 | 0.000 | 0.355 | 0.355 |
| dep ~ |  |  |  |  |  |  |
| idade | -0.002 | 0.001 | -3.036 | 0.002 | -0.002 | -0.033 |
| imc | -0.001 | 0.000 | -2.208 | 0.027 | -0.001 | -0.010 |

Covariances:
|  | Estimate | Std.Err | z-value | P(>\|z\|) | Std.lv | Std.all |
| --- | --- | --- | --- | --- | --- | --- |
| idade ~~ |  |  |  |  |  |  |
| imc | -15.045 | 4.690 | -3.208 | 0.001 | -15.045 | -0.074 |

Intercepts:
|  | Estimate | Std.Err | z-value | P(>\|z\|) | Std.lv | Std.all |
| --- | --- | --- | --- | --- | --- | --- |
| .quant_med_cron | 0.885 | 0.104 | 8.500 | 0.000 | 0.885 | 0.692 |
| .quant_doe_cron | 0.412 | 0.139 | 2.969 | 0.003 | 0.412 | 0.302 |
| .multimorbidade | 0.131 | 0.040 | 3.288 | 0.001 | 0.131 | 0.273 |
| .tempo_s_mdcmnt | 6.145 | 0.126 | 48.878 | 0.000 | 6.145 | 0.837 |
| .depressn_svrty | 0.605 | 0.041 | 14.577 | 0.000 | 0.605 | 0.565 |
| .medic_psi | 0.013 | 0.001 | 9.994 | 0.000 | 0.013 | 0.128 |
| .age_depression | 12.597 | 0.801 | 15.727 | 0.000 | 12.597 | 0.588 |
| idade | 58.270 | 0.155 | 375.310 | 0.000 | 58.270 | 3.798 |
| imc | 27.627 | 0.172 | 160.879 | 0.000 | 27.627 | 2.081 |
| .morbidity | 0.000 |  |  |  | 0.000 | 0.000 |
| .dep | 0.000 |  |  |  | 0.000 | 0.000 |

Variances:
|  | Estimate | Std.Err | z-value | P(>\|z\|) | Std.lv | Std.all |
| --- | --- | --- | --- | --- | --- | --- |
| .morbidity | 1.000 |  |  |  | 0.772 | 0.772 |

|  |  |  |  |  |  |  |
| --- | --- | --- | --- | --- | --- | --- |
| .dep | 1.000 |  |  |  | 0.999 | 0.999 |
| idade | 235.350 | 3.504 | 67.167 | 0.000 | 235.350 | 1.000 |
| imc | 176.177 | 111.421 | 1.581 | 0.114 | 176.177 | 1.000 |
| .quant_med_cron | 0.675 | 0.012 | 54.268 | 0.000 | 0.675 | 0.413 |
| .quant_doe_cron | 0.123 | 0.013 | 9.457 | 0.000 | 0.123 | 0.066 |
| .multimorbidade | 0.091 | 0.001 | 79.826 | 0.000 | 0.091 | 0.391 |
| .tempo_s_mdcmnt | 53.685 | 1.893 | 28.359 | 0.000 | 53.685 | 0.995 |
| .depressn_svrty | 0.274 | 0.020 | 13.408 | 0.000 | 0.274 | 0.239 |
| .medic_psi | 0.009 | 0.001 | 11.540 | 0.000 | 0.009 | 0.962 |
| .age_depression | 128.985 | 7.887 | 16.353 | 0.000 | 128.985 | 0.281 |

